# Development and psychometric evaluation of the Severity of Myalgic Encephalomyelitis Scale (SoMES) and the Myalgic Encephalomyelitis Occupational Status Scale (MEOSS)

**DOI:** 10.64898/2026.11.04.26367265

**Authors:** Sarah F Tyson, Russell Fleming

**Author notes:** Corresponding Author: Prof Sarah Tyson.

## Abstract

**Objective:** To develop and psychometrically evaluate two new brief assessments measuring disease severity (the Severity of ME Scale, SoMES) and occupational status (the ME Occupational Status Scale, MEOSS) in myalgic encephalomyelitis/chronic fatigue syndrome (ME/CFS)

**Methods:** Draft SoMES and MEOSS were co-produced from relevant literature and lived experience. An online survey completed by adults with ME/CFS and was repeated two weeks later to assess reliability. The SF-36 (physical function) assessed criterion-related validity (Spearmann correlations) and discriminant validity (Kruskal-Wallis tests). Weighted kappa co-efficients assessed test-retest reliability and measurement error calculated minimal detectable difference (MDD).

**Results:** 995 adults with ME/CFS participated and 579 completed the retest survey. Construct validity was confirmed from feedback. Criterion related validity was moderate-good (SoMES r = 0.663, MEOSS r=0.472, both p<0.001). Discriminant validity was demonstrated by significant differences in SF36-PF scores in different levels of SoMES and MEOSS (p=0.003 - 0.0001). SoMES’ test-retest reliability was moderate (k =0.53, P<0.001) with a MDD of 1.6, and excellent for the MEOSS (k = 0.76 p<.001) and the MDD was 1.4

**Conclusions:** The Severity of ME Scale (SoMES) and the ME Occupational Status Scale (MEOSS) are valid, reliable, sensitive assessments of ME/CFS severity and occupational status respectively.

## Introduction

Myalgic Encephalomyelitis (ME)/ Chronic Fatigue Syndrome (CFS) is a chronic disease affecting multiple body systems. The hallmark symptoms are debilitating fatigue/exhaustion, post-exertional malaise, cognitive impairment, sleep disturbance and pain while neurological (particularly autonomic), immunological, metabolic and endocrine abnormalities are widely reported^1–3^. It is a highly disabling disease^4,5^. Less than a third of people with ME/CFS can work^6^ and a quarter are severely or very severely affected, leaving them housebound, or even bedbound^7^. Consequently, quality of life for people with ME/ CFS is poor^8^. It is also common, affecting a prevalence of ∼0.6% of the UK population, ie. ∼410,000 people^9^. This is likely an underestimate however, as some 45% of 1.9 million people in the UK with Long COVID are thought to meet the diagnostic criteria for ME/CFS^10^.

Despite its prevalence, impact on quality of life and societal burden^11^, ME/CFS is an under-researched condition. One limiting factor is the lack of psychometrically robust, condition specific patient reported outcome measures (PROMs). Several systematic reviews of PROMs for ME/CFS have concluded that no recommendations could be made^12–16^. Persistent criticisms include the failure of existing measures to capture the issues which are important and relevant to people with ME/CFS (i.e. limited face and content validity), lack of psychometric evaluation, and the use of unvalidated assessments.

The authors sought to address this issue by undertaking a project to co-produce and psychometrically evaluate a series of PROMs to address the difficulties experienced by people with ME/CFS; the ME Association’s Clinical Assessment Toolkit (MEA-CAT). It was designed, primarily for use as a clinical assessment tool, but can also be used for research or service evaluation purposes. It includes PROMs addressing ME/CFS symptoms, post-exertional malaise, activity levels, clinical needs, and satisfaction with clinical services, which are published elsewhere. Here we report the work to develop and evaluate two activity and participation-based assessments which measure ME/CFS severity and occupational status.

Several scales assessing the severity of ME/CFS have been published. They all, in varying degrees describe severity in terms of symptoms, and activity and/or participation limitations. Probably the best-known is the Bell Disability Scale^17^. This is a 0-100 scale, divided into 10 increments with detailed descriptors of symptom severity, functional abilities and work-related limitations defining each level. In 2011, the ME: International Consensus Criteria^18^ produced a shorter, simpler four-level scale which ranged from ‘mild’ (controversially defined as “approximately 50% reduction in activity”) to ‘very severe’ (defined as “bedbound and dependent on help for physical functions”). Another four-increment scale was published in the 2021 NICE Guidelines for ME/CFS^1^. These increments also ranged from ‘mild’ to ‘very severe disability’, but the descriptors were broader and more detailed covering aspects of activity and participation limitations. Most recently, an updated version of the ME Association’s Disability Rating Scale^19^ involves six levels labelled from ‘0% disabled (recovered)’ to ‘90-100% disabled (very severe)’. Detailed descriptions of symptoms, impairments and activity limitations, focussing on mobility and personal care needs define each level. All of these scales appear to be based on the authors’ personal and/or professional experience, we have been unable to find any literature regarding the development or psychometric evaluation of these scales.

A scoping review undertaken in preparation for this project found several studies of occupational status in ME/CFS. A 2004 systematic review of disability, including work status in ME/CFS selected 54 papers found that employment levels were low^20^. However, the conclusions’ strength were limited by the inconsistent information regarding work status. Most merely reported whether participants were employed/ unemployed but these terms were undefined or used inconsistently. For example, it was unclear whether ‘employed’ included part-time as well as full-time work, or whether the ‘unemployed’ were seeking to return to work, were on sick leave, receiving disability related benefits or retired on medical grounds. The authors highlighted the need for a condition-specific validated assessment of occupational capacity. However, no such tool has been produced. Some subsequent publications^21–23^ have attempted to make more nuanced classifications of work status, but these are inconsistent and unvalidated.

As noted above, the aim of this study was to co-produce and psychometrically evaluate two new assessment tools to measure illness severity and occupational status.

## Method

The COSMIN Methodology for Assessing the Content Validity of Patient Reported Outcomes^24^ and the COSMIN Design Checklist for Patient-Reported Outcome Measurement Instruments were used to structure the methodology, analysis and reporting^25^.

### Content Validity

#### Context

This is a patient-led research study. A detailed description of the context for this study has been published previously^26^ and is summarised here. The authors have lived experience of ME/CFS, plus relevant academic and professional backgrounds. They co-produced the PROMs with advisory groups of people with lived experience of ME/CFS and clinicians working in specialist ME/CFS services who provided their expertise at every stage of the project. As detailed in the introduction, the assessments described here formed part of the MEA-CAT and are intended for use in clinical practice or research as an assessment tool, outcome measure, or to monitor change over time.

#### Construct and conceptual framework

Our aim was to produce tools to assess the severity of ME/CFS and occupational status. Following extensive discussion about the construct of ‘ME/CFS severity’ it was decided that this tool should focus on severity in terms of the ‘impact of ME/CFS on daily life’. This was named the Severity of ME Scale (SoMES).

The MEA-CAT includes a detailed assessment of patients’ activity limitations, in terms of what the patient can, or cannot do. This is called the ME Activity Questionnaire (MEAQ) and will be published elsewhere. It does not include ‘work’ as, during the development stages of the MEAQ it was found to be a separate construct to activity limitations and disrupted the scaling properties and construct validity of the MEAQ. However, as most adults with ME/CFS are of working age and work is a high priority for the ME/CFS community, the advisory groups felt it needed to be included in the MEA-CAT, so a new assessment was planned. It needed to encompass ‘work/employment’ in its broadest sense to include paid employment, unpaid work and education. Thus we resolved to refer to ‘occupation’ rather than work, or employment. The assessment, named the ME Occupational Status Scale (MEOSS) also needed to recognise the nuance between ‘paid full employment’ and ‘not working’. It was noted that people with ME/CFS adopted many strategies and adaptations to be ‘occupied’ and these where highly varied. Not only did they depend on the severity of an individual’s illness but also their skills and knowledge, the type of occupation they wanted, and the resources available to them. Thus, we resolved to produce broad categories which accommodated that variability rather than an exhaustive list of the many types and levels of occupation.

#### Target population

Both assessments were intended for use with adults with ME/CFS. Thus, our selection criteria for data collection were broad: Adults with a diagnosis of ME/CFS in the UK.

#### Item generation, response format, and recall period

The existing measures of ME/CFS severity, disability and work status described in the introduction were reviewed by the advisory groups. The following criticisms were made of the existing illness severity scales:

- Members did not understand the concept of ‘% disability’ and could not relate it to their experience, or that of their patients.
- The use of terms ‘very mild/ mild/moderate/ severe /very severe’ were considered misguided. In particular, the terms ‘very mild’ or ‘mild’ were considered misnomers as the degree of activity limitation described, and the impact on everyday life, were far from mild.
- The descriptors:

`o` were too prescriptive and stark. In particular, the symptoms specified did not always match with the activity limitations described. Members felt their illness would often span more than one level.
`o` did not adequately recognise the fluctuating nature of ME/CFS.
`o` gave undue emphasis to physical symptoms and activity limitations. Cognitive limitations were particularly underrepresented.

The assessments of occupational status focussed on paid employment, which was too crude and did not acknowledge the ‘reasonable adjustments’ and other measures people with ME/CFS often made to return to, or stay in employment. They also did not acknowledge the substantial proportion of the ME community who were engaged in some form of education, or who undertook unpaid work. Consequently, the SoMES and the MEOSS were constructed with a simple, single question checklist format. Participants merely identified which category best described their situation. In keeping with the other assessments in the MEA-CAT, the recall period was one month.

For the SoMES, neutral terms (levels 1-5) were used to define categories, rather than percentages or labels such as mild/moderate/severe. The descriptors of each category focussed on the impact of the ME/CFS on daily life in terms of the activities that users could undertake and the adaptations and/or assistance needed. We used general, neutral, flexible language, minimised mention of specific symptoms and activities, and highlighted the spectrum within levels. Intermittent levels (e.g. between levels 1 and 2) were added to accommodate people whose abilities fell, or fluctuated between levels. However, these were given the same score as the preceding level. A note was added to emphasise that ‘activities’ could be physical, cognitive, social, or emotional etc.

For the MEOSS, advisory group members felt it important to highlight the intermittent stages between ‘employed’ and ‘unemployed’, and to recognise unpaid work and education. These were categorised into 5 levels (scoring from 0-4), which covered: not working; on sick (or other) leave; unpaid work; working with accommodations; and working without adjustments for ME/CFS. Each category had a more detailed description. A supplementary question about job instability and the difficulties the patient was facing was added for use in clinical practice. This was not scored.

#### Construct validity: Evaluation of relevance, comprehensibility and comprehensiveness

The protype SoMES and MEOSS was devised and completed by the advisory group members, who then completed a cognitive debriefing interview regarding

- comprehensibility of the levels and descriptors
- relevance of the levels and descriptors
- whether there was sufficient separation between the levels
- whether there was any duplication between levels
- their thoughts in using intermediate levels in the MEOSS (e.g. ‘between levels 2 and 3’)
- comprehensiveness and whether anything should be added
- ease of completion
- whether any other changes were needed.

These interviews were completed by email as this was the most accessible form of communication for the group members. The interview guide, with prompts was emailed to them with further discussion (by email, phone or video call as preferred) as necessary. As the information sought was quite specific, deductive framework analysis was used to analyse the advisory groups’ response using the broad codes of relevance; duplication; comprehensiveness; comprehensibility/ease of use; and accommodations. Internal member checking ensured representativeness/internal validity. As a final triangulation, participants in the online survey (details below) were invited to add feedback regarding the SoMES and MEOSS in an open question asking if there was anything they would like to tell us about the scales and how they might be improved. These comments were analysed using the same deductive framework.

### Criterion-related validity, discriminant validity, test-retest reliability and minimal detectable difference (MDD)

A classic theory testing approach^25^ was used to assess the other aspects of validity and reliability. The prototype SoMES and MEOSS were converted to an online survey using the Qualtrics Survey tool. Adults in the UK who had been diagnosed with ME/CFS were invited to complete them online. Recruitment was via publicity in the ME Association’s social media and publicity channels, ME/CFS support groups and the 25% ME group (a charity for people with severe ME/CFS). In addition to the SoMES and MEOSS, participants completed the physical function section of the Medical Outcomes Scale Short form (SF36-PF)^27^. Spearman correlations assessed criterion-related validity, testing the hypothesis that higher physical-related quality of life (SF36-PF scores) would be related to ME/CFS severity (higher SoMES scores) and occupational status (higher MEOSS scores). Kruskal-Wallis tests assessed divergent validity by testing whether there were significant differences in SF-36PF scores between different levels of the SoMES and MEOSS.

To assess test-retest reliability and minimal detectable difference, participants were asked to repeat the survey two weeks after submission. In addition to completing the SoMES and MEOSS, participants were asked whether ‘overall’, their ME/CFS had changed over the previous two weeks. The data from those who responded that their ME/CFS was ‘the same’ were used in the analysis. Weighted kappa scores assessed test-retest reliability, while the MDD was calculated from the measurement error where:

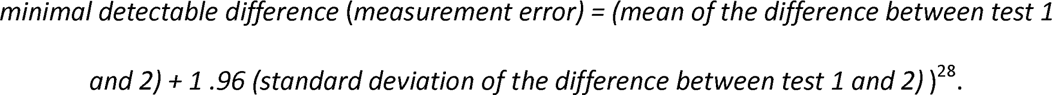

## Results

### Content validity

At the cognitive debrief of the prototype SoMES and MEOSS, the advisory group members agreed that the assessments were comprehensive, relevant, easy to understand and complete, and the levels and descriptors reflected their experience. This was confirmed in the feedback from survey participants.

Nine-hundred and ninety-five participants completed the online survey and 579 completed the retest survey. Their details are shown in Table 1. Three hundred and fifty-four (61.1%) participants reported their ME/CFS was unchanged since the first survey, 46 (7.9%) had improved and 179 (30.9%) had deteriorated.

**Table 1.** showing the participants’ characteristics.

|  | First test n= 995 | Retest n= 579 |
|---|---|---|
| Gender | Women (n=832, 83.6%)<br>Men (n=146, 14.7%)<br>Other (15, 1.5%) | Women (n=484, 83.6%)<br>Men (n= 85, 14.7%)<br>Other (n= 10, 1.6%) |
| Mean age (years) | 54.14<br>(sd 18.79, range 17-71) | 53.78<br>(sd 13.21, range 18-83) |
| Mean time since diagnosis (years) | 12.80<br>(sd 9.0, range 0.25-50). | 12.67<br>(sd 10.63, range 0.5- 50) |
| Mean SF36-PF score (max 100) | 25.7 (sd 21.55 range 0-95). | 25.95 (sd 21.12, range 0-95) |
| SoMES score | 2.87/ 5 (sd 0.88)<br>Median 3.0 (IRQ 2-4) | Mean 2.96 (sd 0.91)<br>Median 3.0 (IRQ 2-4) |
| Adapting activity:<br>level 1 and 1-2 | 51 (5.1%) | 27 (4.8%) |
| Prioritising activities:<br>level 2 and 2-3 | 284 (28.6%) | 158 (27.2%) |
| Limited independence:<br>level 3 and 3-4 | 399 (40.1%) | 215 (37.2%) |
| Essentially housebound:<br>level 4 and 4-5 | 227 (22.8%) | 169 (29.2%) |
| Essentially bedbound:<br>level 5 | 15 (1.5%) | 9 (1.6%) |
| MEOSS score | Mean 0.95 (sd 1.35)<br>Median 0.00 (IRQ 0,3) | Mean 0.87 (sd 1.33)<br>Median 0.00 (IRQ 0,2) |
| Not working/ studying | 630 (64.7%) | 388 (67.1%) |
| On sick leave (or other type of leave) | 63 (6.5%) | 36 (6.2%) |
| Unpaid work/ informal study | 21 (2.2%) | 12 (2.1%) |
| Working/ studying with accommodations | 241 (24.8%) | 128 (22.1%) |
| Working/ studying without<br>adjustments for ME/CFS | 18 (1.8%) | 14 (2.4%) |

The correlation between the SF36-PF and the SoMES was -0.663 (p<0.001) indicating strong criterion-related validity. Kruskal-Wallis tests indicated highly significant differences in SF36-PF scores between levels of the SoMES showed strong divergent validity. This was both overall and in each pairwise comparison (p=0.003 - 0.0001) except the comparison between levels 4 and 5. This was likely underpowered as there were only 15 participants at level 5. Test-retest reliability was good (k =0.53, P<0.001) and the minimal detectable difference was 1.6.

The correlation between the SF36-PF and the MEOSS was 0.472 (p<0.001) indicating moderate, but highly significant criterion-related validity. Kruskal-Wallis tests showed strong divergent validity in that there were highly significant differences in SF36-PF scores between levels of the MEOSS (p<0.01). However, only the individual comparisons between ‘not working/studying’ and ‘working/studying with and without accommodations’ were significant (p = 0.001 and 0.004 respectively). Test-retest reliability was excellent (k = 0.76 p<.001) and the minimal detectable difference was 1.4

## Discussion

The results of this study show the SoMES and MEOSS are valid, reliable, sensitive measures of ME/CFS severity and occupational status respectively. As noted in the introduction, there are no previous reports of the psychometric properties of measurement tools for these constructs so comparisons with other reports are not possible. The main strength of this study lies in its large sample size and co-production. By using a co-production approach with people with ME/CFS and clinicians in ME/CFS specialist services, we are confident they represent the issues that are important and relevant to people with ME/CFS and clinical services in an accessible format.

The main limitation lies in the representativeness of the sample. Like other large convenience surveys of people with ME/CFS^9,29,30^, our sample was predominantly middle-aged women with extensive lived experience who are moderately affected. However, the recruitment methods may have biased towards people who are active on social media, and in ME charities and other organisations. There is also evidence^21,23,31–34^ that samples recruited from ME/CFS services, or community- or registry-based surveys tend to be younger, have shorter disease duration, be less severely affected, and a greater proportion who are working or intending to return to work, than those who respond to surveys. However, whether these differences would impact on validity or reliability of the SoMES and MEOSS is moot.

## Conclusions

The Severity of ME Scale and the ME Occupational Status Scale e are valid and reliable measures of ME/CFS Severity (in terms of the impact on everyday life) and occupational status in ME/CFS. Copies are found in the supplementary files or the ME Association website (<u>The ME Association Clinical Assessment Toolkit (MEA-CAT) - The ME Association</u>) and are freely available for others to use.

## Data Availability

Data produced in the present study are available upon reasonable request to the authors

## Acknowledgments

The authors would like to thank the ME advisory group and clinicians’ advisory groups who co-produced this work.

## Supplementary File 1. The Severity of ME Scale (SoMES)

Please choose which level best describes how ME/CFS affects you ‘in the last month’:

PLEASE NOTE: Activities can be physical, cognitive, social, emotional etc.

- <u>Level 1: Adapting activity</u>. You can do the things that are important to you, but may need to adapt how much you do, or how you do them.
- <u>Level 1-2.</u> In between levels 1 and 2.
- <u>Level 2: Prioritising Activities.</u> You can do the things that are most important to you, but you limit or avoid other activities to prioritise them. For example: stopping hobbies to prioritise work (or vice versa)
- <u>Level 2-3.</u> In between levels 2 and 3.
- <u>Level 3: Limited independence.</u> You can carry out some light/ non-strenuous activities but with rests. You will need help with some activities (including having someone else to do them) e.g. domestic chores.
- <u>Level 3-4.</u> In between levels 3 and 4.
- <u>Level 4: Essentially housebound</u>. You need help with many activities such as domestic chores or personal care (including having someone else to do the chores). Mobility is probably restricted so that it is challenging to travel outside your home. You may need a wheelchair (or similar) and/or to travel by a vehicle (eg car) if going outside.
- <u>Level 4-5</u>. In between levels 4 and 5.
- <u>Level 5: Essentially bedbound</u>. You will need help or someone else to complete most activities for you. At the most severe end of this level, this <u>could</u> include difficulty eating and/or the need to be artificially fed, environmental adaptations to accommodate hypersensitivity to sound, light and/or touch, or support for severe cognitive impairments. Mobility is probably severely restricted and you will spend most of you time lying down e.g. in bed or the settee.

### ME Occupational Status Scale

NOTE This refers to both paid and unpaid work or education. What has been your work situation over the last month?

1. I am not seeking employment (e.g. retired on age or health grounds, on long term benefits, homemaker) (Score 0)
2. I am on sick leave (or other type of leave) and would like to return to work /study. (Score 1)
3. I do flexible, unpaid work / studying (e.g. volunteering/ informal education) (Score 2)
4. I am working/ studying, but with accommodations (e.g. extra equipment; flexible hours; ‘light duties’; extra time) OR with changes to my job to reduce demands (e.g. self-employed, part time, less demanding role) (Score 3)
5. I am working/ studying without any accommodations for my ME/CFS. (Score 4)

NB If the MEOSS is being used for clinical purposes, add the following question

If you are working/studying, are you concerned about being able to keep your job or complete your course? Answer Yes/no (but do not include in the score)?

If yes, please give details, (Free text box)

